# Sotalol dose optimization for fetal tachycardia: a pregnancy physiologically based pharmacokinetic model study

**DOI:** 10.1101/2024.12.17.24319139

**Authors:** Hedwig van Hove, Joyce E.M. van der Heijden, Anne van Uden, Violette M.G.J. Gijsen, Rick Greupink, Saskia N. de Wildt, Joris van Drongelen

## Abstract

**Objective:** To establish an optimized sotalol dosing strategy for fetal tachycardia by using a pregnancy computational model for dose simulations.

**Methods:** A physiologically-based computational model, including pregnancy-related changes and placental transfer values, was established and verified. Simulations of the current dosing advises and prospective dosing scenarios were performed. To avoid maternal dose-related toxicity (QT-prolongation) we aimed for maternal concentrations <2.5 mg/L. Based on neonatal concentration-effect data, we aimed for a fetal C_trough_ concentrations of 0.4 – 1.0 mg/L.

**Results:** The pregnancy physiologically-based pharmacokinetic model accurately predicted maternal and fetal exposures. Predictions indicate that almost 16% of maternal plasma concentrations exceed the toxic level of 2.5 mg/L at the maximum oral daily dose of 480 milligram, while 90% of fetuses have a C_trough_ concentration within the therapeutic window. When lowering the maximum daily dose to 400 mg, 0.1% of maternal plasma concentrations exceed 2.5 mg/L, while 87% of the fetal plasma concentrations remain in the therapeutic window. Additionally dosing 480 mg in three times daily reduces the risk of high maternal plasma exposure to 0.3%, while maintaining effective fetal C_trough_ concentrations in 95% of fetuses.

**Conclusion:** Pregnancy computational modeling can be used to adequately predict maternal and fetal sotalol exposures. Our simulations suggest that daily doses should not exceed 400 milligram and that dividing the oral daily dose over three doses improves the balance between high maternal plasma exposure and effective fetal concentrations.

**Funding:** This publication is based on research funded by the Bill & Melinda Gates Foundation (INV-023795).

## Introduction

Fetal arrhythmias occur in about 1% of pregnancies and sotalol, a beta-blocker, is one of the proposed first choice therapeutic options (1, 2). The variety of dosing regimens of sotalol for fetal tachycardia is based on scarce evidence. Only few studies have been performed, with relatively small numbers of participants, but all studies show that only part of the group receiving sotalol is responsive to the therapy (3–6). Next to this, clinical evidence shows a high incidence of maternal side effects related to sotalol use for transplacental therapy (7, 8).

Pharmacotherapy during pregnancy is challenging due to potential teratogenicity, but also due to the physiological changes that occur during pregnancy which impact drug disposition, e.g. increased glomerular filtration rate (GFR) and changes in drug metabolizing enzymes (9). These changes are more profound later in pregnancy and are suggested to contribute to more rapid clearance of sotalol in pregnancy, since sotalol is predominantly cleared renally (10, 11). The emergence of computational models, simulating how pregnancy-related changes in women’s bodies affect required drug doses for mothers and children throughout pregnancy, can provide a solution to clinical issues surrounding correct doses of medicines. These models, also known as physiologically based pharmacokinetic (PBPK) models, act as predictive tools to investigate drug levels in the mother and fetus upon varying administrations. These predictions can support the medicine benefit–risk decision and inform dose adjustment in a pregnant population (12).

Therefore, we aimed to evaluate the current sotalol dosing strategy by developing a pregnancy PBPK model that accurately describes maternal and fetal exposure. Next, we aimed to evaluate exposure with current dosing and to suggest an optimized dosing strategy in which a better balance between fetal efficacy and maternal safety is reached.

## Methods

Briefly, in this study a pregnancy PBPK model was extended with Caco-_2_ apparent permeability values to represent placental transfer parameters. This allowed the prediction of both maternal and fetal exposure. Model performance was first verified against published maternal and fetal pharmacokinetic data. Next, alternative sotalol dosing regimens were simulated to establish a model-informed dose aiming at a fetal C_trough_ concentration of 0.4-1.0 mg/L while minimizing maternal peak concentrations.

All simulations were conducted in Simcyp v21 (Certara UK Limited, Simcyp Division, Sheffield, UK). Although the sotalol compound model was available in the Simcyp repository, some adjustments were necessary to enable accurate predictions of oral dosing (13). The advanced dissolution, absorption and metabolism (ADAM) model implemented within Simcyp was used to optimize oral absorption predictions of sotalol. The optimized compound model can be found in the Supporting Information, Table S1. We mined sotalol clinical data from PubMed for healthy non-pregnant and pregnant individuals (Table S2 and S3). Observed data are then compared to predicted values to verify the sotalol compound model and the performance of the healthy adult and pregnant population predictions of the PBPK model. The pregnant physiology model was directly available in the Simcyp software (i.e., Sim-Pregnant population). To assess whether gestational changes influence sotalol exposure during pregnancy, simulations including different gestational age (GA) groups were executed (i.e. GA of 24, 32 and 40 weeks).

### 2.1 Apparent permeability of sotalol

The apparent permeability coefficient (P_app_) represents the permeability of sotalol over a Caco-2 cell monolayer. Caco-2 cell assay data is commonly used for estimation of pharmaceuticals across barrier tissues and was now used as input for transplacental transfer in the permeability-limited placental model in Simcyp (14, 15). Previously determined P_app_ values were used (16). For the exact calculations, see Supporting Information.

### 2.2 Predictive performance PBPK model

Predictive performance of both the healthy volunteer and the pregnant women PBPK model was evaluated by a visual predictive check and by calculating the ratio of predicted-to-observed PK parameters. For a visual check, observed plasma concentration-time profiles were extracted from literature, digitalized with WebPlotDigitizer v4.6. Predicted plasma concentration-time curves were compared to the observed values. Predicted-to-observed (p/o) ratios were calculated and ratios within 0.5 to 2-fold range were considered acceptable (17).

### 2.3 Current sotalol dosing advise

Most guidelines propose starting doses of 160 mg to 320 mg, given orally divided in two to three doses a day. If the desired effect has not been achieved after three days of therapy, the dose will be increased by 80 mg/day with a maximum daily dose of 480 mg/day (3, 4, 18). The therapy is judged ineffective if the fetal arrhythmia does not resolves or improves (e.g. heart rhythm decreases below 200 beats per minute) three or more days after the start of the highest sotalol dose (19). In that case, a combination therapy including digoxin or flecainide is advised (20, 21).

### 2.4 Pregnancy PBPK modeling

For all simulations, i.e. verification of prospective, the default pregnancy physiology model was used, incorporating gestational age related changes in GFR that affect sotalol clearance (22). To predict fetal exposure, the sotalol P_app_ values were included for placental transfer. Simulations of current dosing advise scenarios were conducted to determine whether the fetal target concentration was achieved without surpassing the maternal toxic target. Last, we included prospective simulations with alternative dosing scenarios.

### 2.5 Maternal and fetal exposure targets

To allow re-evaluation of the current dosing regimen of sotalol during pregnancy to treat fetal tachycardia, clear target concentrations need to be maintained. The study by Oudijk et al. 2003 suggests that all maternal serum levels should remain below the level of 2.5 mg/L(18). Next to this, literature data and in-house clinical data (V. Gijsen, unpublished observations) show maternal side effects in up to 70% of cases, even at low doses. A pediatric PK/PD modeling study determined a therapeutic window for children with supraventricular tachycardia (SVT), including neonates, based on comprehensive *in silico* predictions, of a plasma trough concentration (C_trough_) of 0.4 - 1.0 mg/L (23). We therefore applied this therapeutic window for C_trough_ concentrations between 0.4 and 1.0 mg/L in fetal plasma concentration predictions to assess successful fetal therapy.

## Results

### 3.1 Healthy volunteer PBPK model verification

To assess predictive accuracy of sotalol exposure in healthy volunteers, visual predictive checks were conducted for IV (Supporting Information, Figure S1) or PO administration (Supporting Information, Figure S2). Additionally, predicted-to-observed PK parameter ratios were calculated to verify model performance (Figure 1A and B). Observed data are obtained from different pharmacokinetic studies (24, 25). Overall, the PBPK model was considered adequate to predict PK in healthy volunteers.

**Figure 1.**
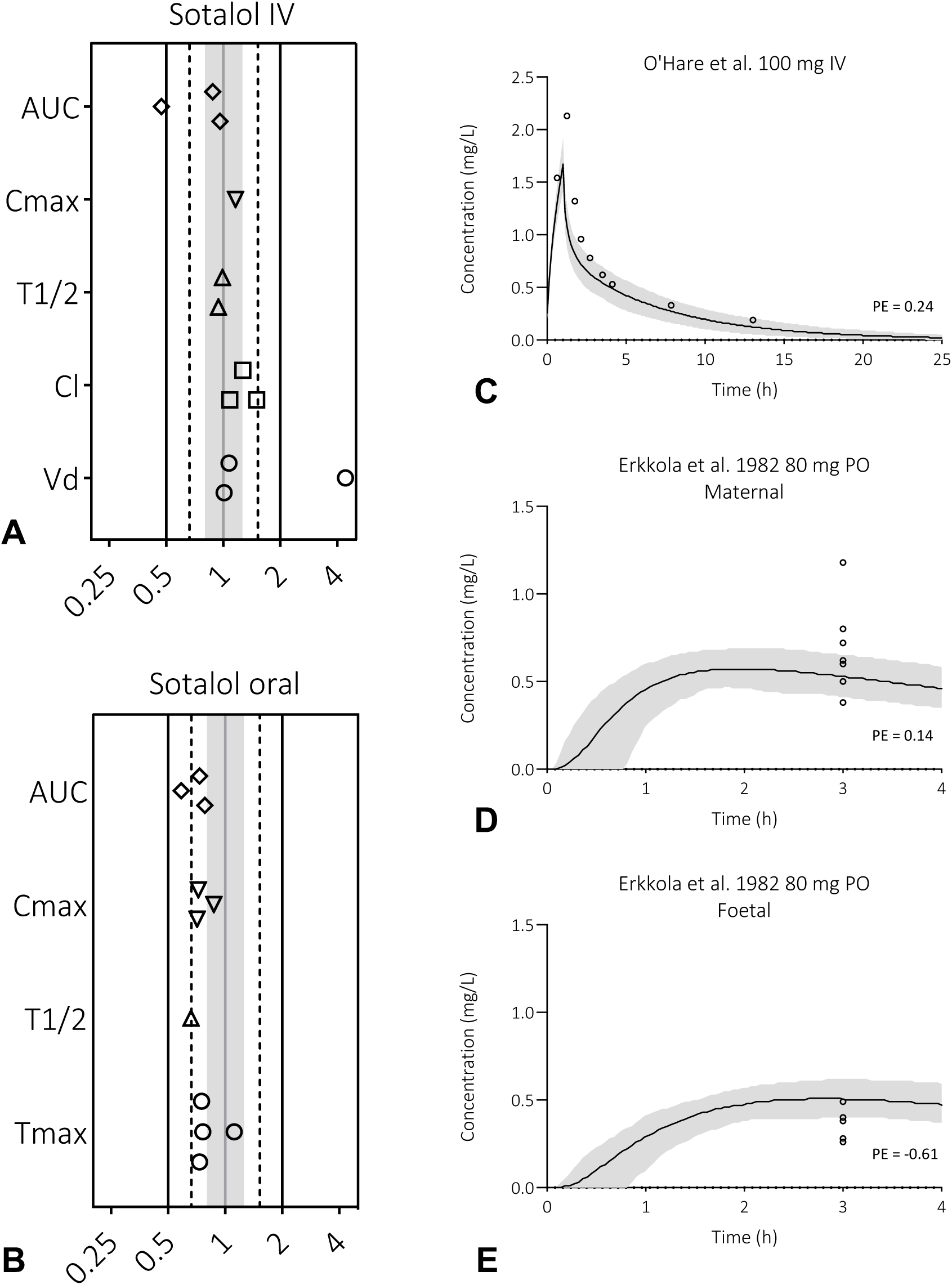
Model performance in relation to reported sotalol plasma concentrations in healthy volunteers, pregnant women and their unborn children. Predicted-to-observed ratios of the area under the curve (AUC) and the maximum concentration (C_max_) for sotalol for healthy volunteers, separated for intravenously (1A) or oral (1B) administration route. Single symbols represent a predicted-to-observed ratio of the average reported parameters for an individual pharmacokinetic study. The black lines represent the 2-fold range, the dashed lines the 1.5-fold range, the grey shaded area represent the 1.25-fold range and the grey line represents the unity line. Observed data are obtained from different pharmacokinetic studies [24, 25]. The prediction of sotalol plasma concentration-time profiles in pregnant women and their unborn children is shown in 1C, 1D and 1E. Here, the solid line is the predicted mean of the simulated population and the shaded area represents the 5^th^ to 95^th^ percentile of the virtual population. Open circles are the observed data, for an IV study (C, data points represent represents the mean [10]) and an oral study (D, individual data points [26]). For the oral study, a simulation of umbilical cord blood concentrations and matching observed clinical data are provided as well (E, individual data points). Abbreviations: oral (PO), intravenously (IV), milligram (mg).

### 3.2 Pregnancy PBPK model verification

Next, the model was applied to the Simcyp default pregnant population to evaluate sotalol exposure (upon IV and PO administration) in pregnant women. Simulated plasma concentration-time profiles of women receiving a 100 mg IV dose or a 80 mg oral dose are shown in Figure 1C and D. Fetal exposure after 80 mg oral dosing is predicted as well and included in Figure 1E. More simulated plasma concentration-time profiles of pregnant women and their unborn children are included in the Supporting Information, Figure S3, S4 and S5. Only two area under the curve (AUC) values were reported for dosing during pregnancy (i.e. one AUC for IV and one for PO dosing) with p/o ratios of 0.71 (IV) and 0.80 (oral), respectively. Taking all together, model performance was considered adequate to predict PK in pregnant women and their unborn children.

### 3.3 Current dosing advise

To determine whether a difference in exposure can be expected at different gestational ages, simulations with the highest recommended oral dose of 240 mg twice daily (BID) were performed in pregnant virtual populations with a gestational age of 24 weeks, 32 weeks and 40 weeks. Figure 2 shows visually no deviation in exposure between the three groups, so maternal and fetal exposure during current dosing regimen will only be performed using a gestational age of 32 weeks, since this GA is most commonly seen in clinical practice.

**Figure 2.**
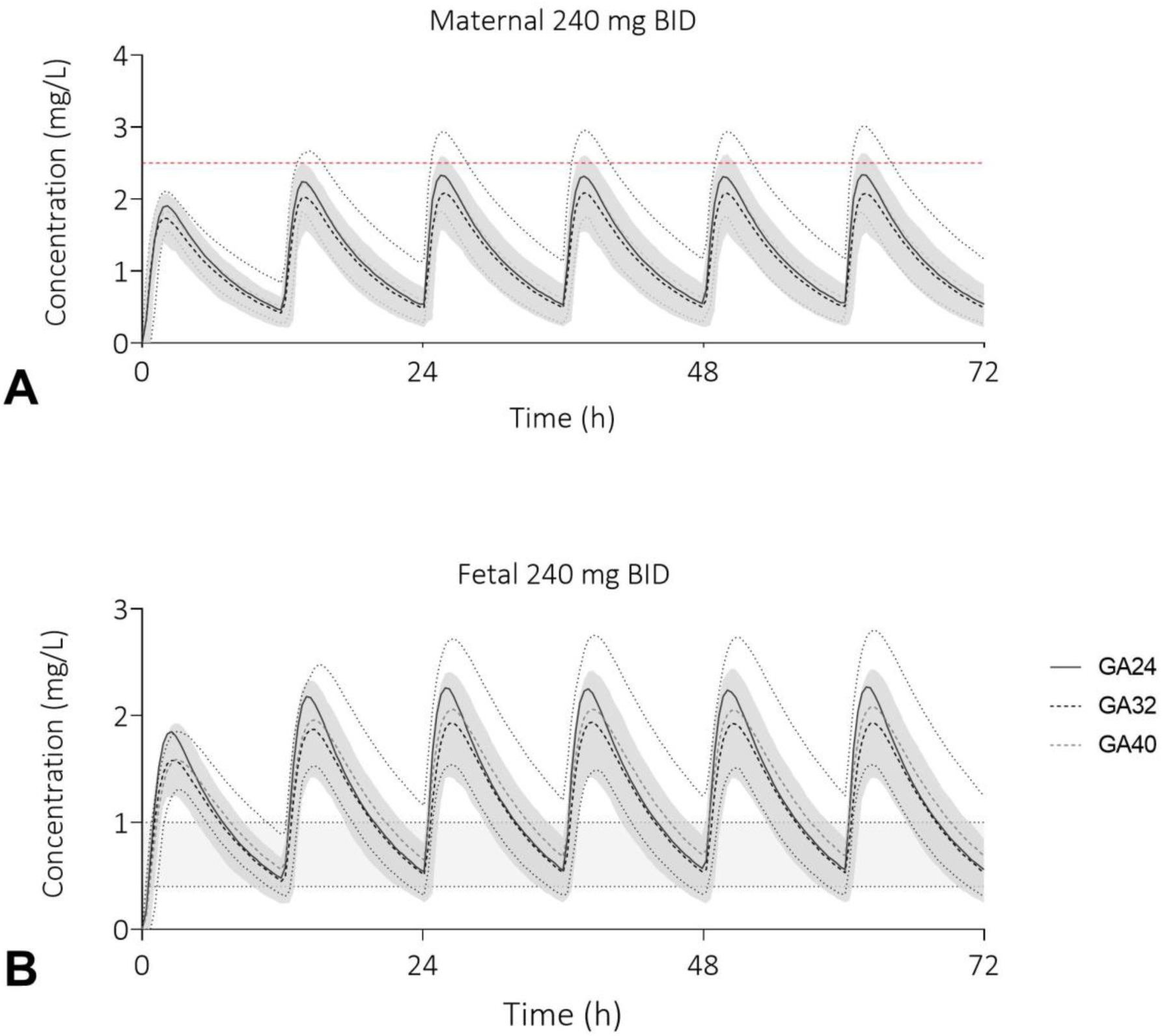
Predicted maternal and fetal sotalol plasma concentration-time profiles at different gestational ages. The shaded area represents the 5^th^ to 95^th^ percentile of the virtual populations, the red dotted line (A) the toxic level of 2.5 mg/L and the grey area (B) the fetal therapeutic C_trough_ window of 0.4-1.0 mg/L. Abbreviations: gestational age in weeks (GA), twice daily (BID), oral (PO), milligram (mg).

To provide insight in maternal and fetal exposure during sotalol therapy to treat fetal tachycardia, the current dosing advise of starting with 80 mg BID and uptitrating to 240 mg BID was simulated. Each dose was given orally twice daily in a fasted state, and therapy was continued for 72 hours. The starting dose was 160 mg/day, which was increased in steps of 80 mg/day (Figure S6 and Figure S7). The mean predicted maternal and fetal plasma concentrations after a daily dose of 400 mg/day and 480 mg/day are presented in Figure 3. 15.6% of the maternal plasma concentrations are predicted to exceed the toxic plasma concentration of 2.5 mg/L at a daily dose of 480 mg, while 89.7% of all fetal C_trough_ concentrations fall within the therapeutic window of 0.4-1.0 mg/L. After a daily dose of 400 mg only 0.1% of maternal subjects is predicted to exceed the toxic level, while 86.2% of fetal subjects is predicted to have fetal C_trough_ concentrations within the therapeutic window (Figure 3B).

**Figure 3.**
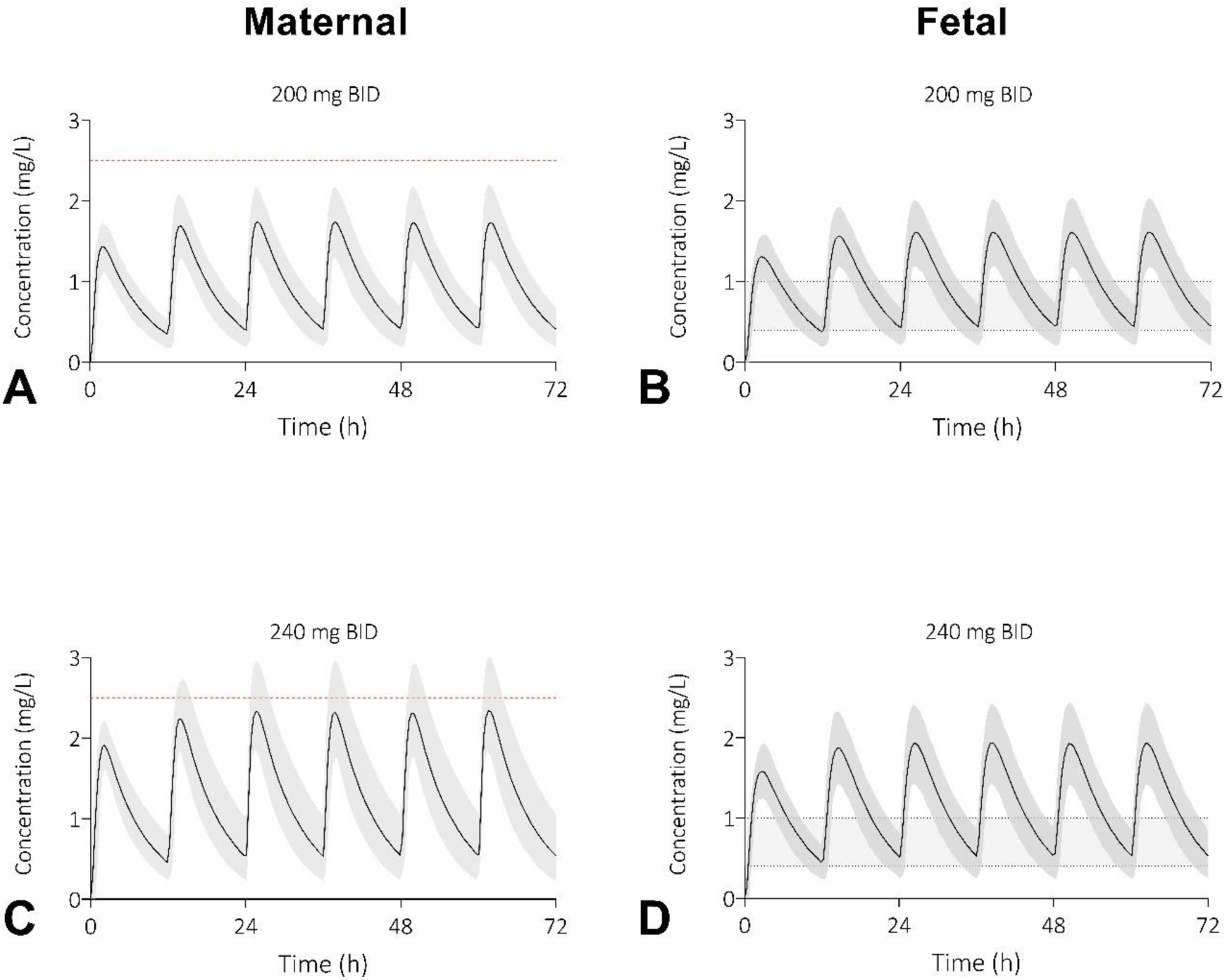
Predicted plasma concentration-time profiles of sotalol for pregnant women and their unborn children after an oral dose of 400 mg/day (A & B) or 480 mg/day (C & D) orally. The shaded area represents the 5^th^ to 95^th^ percentile of the virtual populations, the red dotted line the toxic level of 2.5 mg/L and the grey area the fetal therapeutic C_trough_ window of 0.4-1.0 mg/L (B&D). Abbreviations: twice daily (BID), milligram (mg).

### 3.4 Prospective simulation

Alternative dosing scenarios, in which sotalol was prescribed orally three times a day (TID) were explored for 160, 120 and 80 mg, respectively (Figure 4). When the current maximum dose is maintained, but 160 mg TID, only 0.3% of the maternal plasma concentrations are predicted to exceed the toxic level of 2.5 mg/L, while 98.4 % of fetuses have a C_trough_ between 0.4-1.0 mg/L. For 120 and 80 mg TID none of the predicted plasma concentration are higher than 2.5 mg/L. 95% of fetuses have a C_trough_ between 0.4 – 1.0 mg/L when prescribed 120 mg TID, and for 80mg TID this accounts for 79.0% of fetuses. Fetal plasma exposure after 40 mg TID was assessed as insufficient (Figure S7).

**Figure 4.**
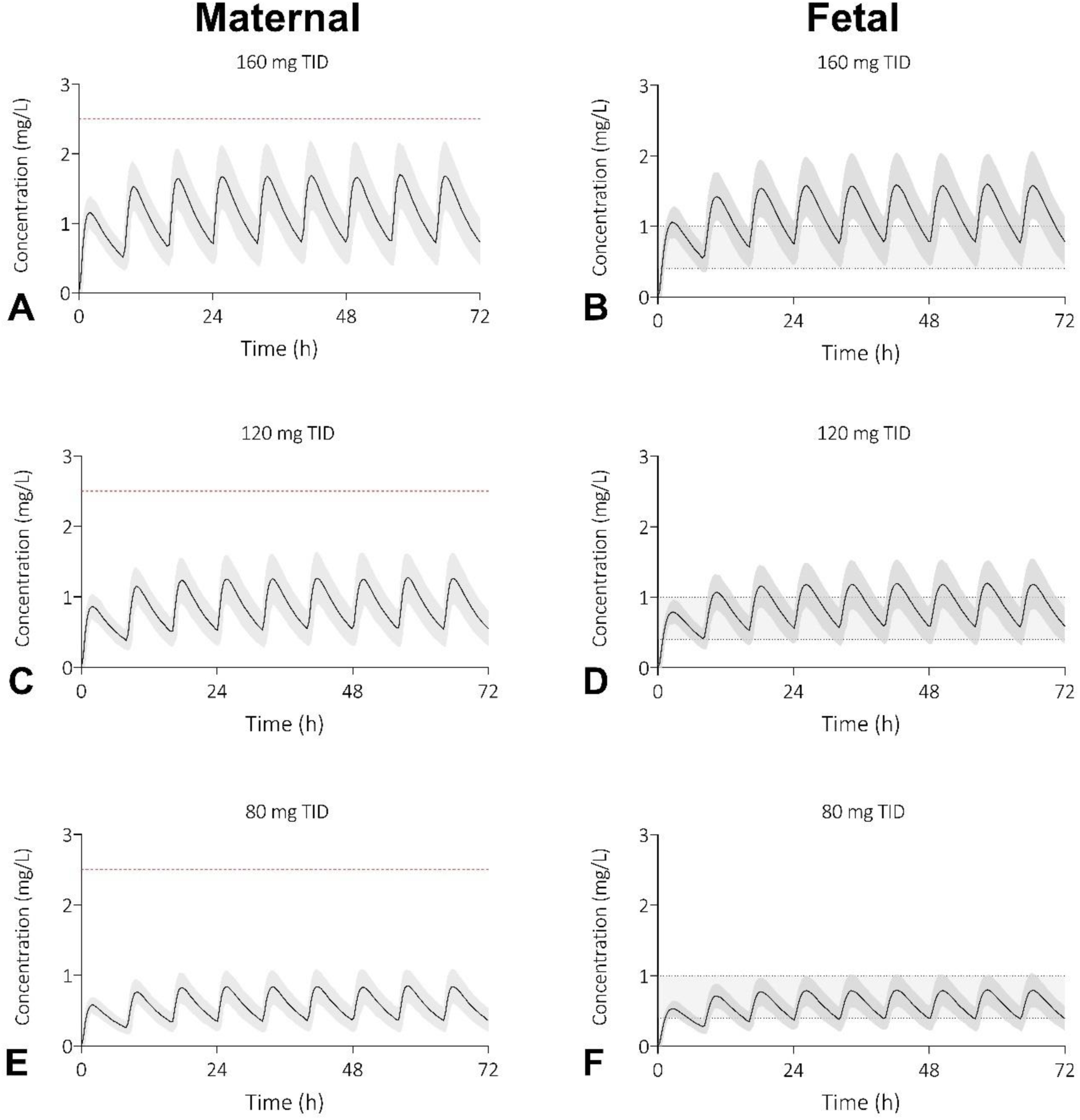
Prospective simulations of sotalol plasma concentration-time profiles for pregnant women and their unborn children after an oral dose of 480 mg/day (A & B), 360 mg/day (C & D) or 240 mg/day (E&F) orally. The shaded area represents the 5^th^ to 95^th^ percentile of the virtual populations, the red dotted line the toxic level of 2.5 mg/L and the grey area the fetal therapeutic C_trough_ window of 0.4-1.0 mg/L (B&D). Abbreviations: three times a day (TID), milligram (mg).

## Discussion

### Main Findings

In this study, a combined approach of pregnancy computational (PBPK) modeling and experimentally established placental transfer of sotalol was applied. The model accurately predicted healthy non-pregnant volunteer, maternal and fetal exposures to sotalol. In pregnancy, finding the therapeutic optimum is challenging, as the benefits and side effects for both the mother and the fetus must be balanced. We present evidence that the current highest recommended dose of 240 mg twice a day (480 mg daily) may contribute less to a positive fetal and maternal outcome than previously thought. At 200 mg twice daily (400 mg daily) sotalol concentrations are already within therapeutic range in 86% of fetuses, with a toxic level in 0.1% of mothers. Increasing the dose to 240 mg twice daily (480 mg daily) leads to an additional 4% of fetus within the therapeutic range, but with maternal concentration above the toxic level in 16%. In addition, spreading the 480 mg daily dose over three instead of two doses seems to be the best, as 98% of fetus are within the therapeutic range with a 0.3% of the maternal concentrations above the toxic level.

The predicted fraction of mothers exceeding the toxic plasma concentration level of 2.5 mg/L after receiving the current highest recommended dose of 240 mg twice a day (480 mg daily) is disproportional (16%) to the predicted fraction of unborn children who benefit from the treatment trough therapeutic levels as compared to 200 mg twice a day (90% vs. 87%). These predictions reflect literature findings of up to 70 to 80% maternal side effect incidence with sotalol use during pregnancy (3, 4, 19). This implies the necessity for a new sotalol dosing strategy, since further increasing the dose beyond 400 mg daily seems pointless.

### Strengths and Limitations

Studying *in vivo* drug exposure in the fetus during gestation is impossible without exposing the pregnancy to risks. PBPK modeling can be a good alternative, since the model is able to mimic *in vivo* scenarios including biological variation between subjects, but predictions need to be reliable. Therefore, assumption in the PBPK model need to be based on the best available evidence. We based the placental passage on outcomes of an *in vitro* Caco-2 model. The Caco-2 cell monolayer acts as surrogate for the placental barrier, enabling transfer studies representative for placental passage. This approach has already been successfully applied in the past (26). Outcomes of the sotalol Caco-2 study were comparable to several peripartum studies reporting extensive passage of the drug over the placental barrier, with a maternal to cord blood ratio of about 1 (18, 27, 28). Additionally, model performance was increased by using the ‘redefine subjects over time option’ for simulations exceeding seven days duration. This ensures that virtual subjects undergo anatomical and physiological changes in line with those observed in clinical practice, enabling more accurate predictions (e.g. increase in renal clearance during pregnancy) (22). These adaptations resulted in accurate estimations of fetal exposure when compared to data from peripartum studies (3–6). Last, model confidence was increased by verifying the model against maternal and fetal clinical datasets derived from different dosing regimens, including multiple dosing settings.

The fetal target window for the C_trough_ is based on a well validated pharmacokinetic-pharmacodynamic model in neonates and older children for SVT. It is assumed that a C_trough_ of 0.4 to 1.0 mg/L should, in addition to the populations described in the paper of Läer et al., also be sufficient in the treatment of fetal tachycardia (23). However, there remains some uncertainty as this model did not take into account age-dependent sensitivity toward pharmacologic and toxicologic effects to sotalol for fetuses. In reality, it might be that even a lower C_trough_ is efficient in a fetal population.

### Interpretation

Although severe fetal tachyarrhythmia develops in <0.1% of pregnancies, the importance of timely and effective treatment of fetal tachycardia during pregnancy cannot be overstated, as it directly influences the cardiovascular development and overall health of the fetus (19). If untreated, fetal tachycardia is associated with fetal hydrops, neurological morbidity and intrauterine death. Best treatment options and dosing strategies would preferably be investigated in a clinical trial setting, however, this is very time-consuming and costly research to perform. PBPK modeling is a modern technique that can support developing optimal dosing regimens by mimicking the *in vivo* situation. To overcome high maternal plasma exposure, while maintaining effective fetal C_trough_ concentrations, our PBPK analyses propose to consider dividing oral daily doses of 480 mg over three doses of 160 mg (Figure 4). This strategy seems adequate to treat most fetuses (98%) successfully and reducing the maternal side-effects to 0.3%. To our knowledge there is no direct literature available on this drug strategy. It would be worthwhile to investigate whether predicted exposures in this study align with clinical practice.

In line with previous reported clinical observations, part of the predicted fetal values do not reach the desired C_trough_ concentrations during therapy. This might explain the absence of fetal response sometimes seen in clinical practice (29, 30). Current advises already propose the use of combination therapy when fetal response is absent (5, 19). Numerous retrospective studies have focused on prenatal diagnosis and management, using sotalol, digoxin, and/or flecainide either as individual agents or in combined therapeutic regimens (4, 31, 32). However, there is no consensus on a superior first-line drug to treat fetal tachycardia. The FAST trial might provide the answer, however, outcomes are not reported yet (33).

What became evident during our study is that the frequency and the extend of maternal side effects, in relation to administered dose, is often reported poorly. Also, reports often lack dosing information. This type of information is important for evaluating quality of care and dosing strategies and should therefore be reported more transparently.

## Conclusion

In conclusion, we demonstrated that a combined approach of pregnancy PBPK modeling and experimentally established placental transfer of sotalol can be informative for clinical dosing. The predictions showed that current dosing regimen in pregnant women should be optimized to prevent maternal side effects. We here suggest another drug or drug-combination if fetal response is not forthcoming at a daily dose of 400 mg/day. Next to this, we propose to consider dividing oral daily doses of 240 mg over three doses to adequately prevent high maternal plasma exposure, while maintaining effective fetal C_trough_ concentrations.

## Data Availability

All data produced in the present study are available upon reasonable request to the authors

## Acknowledgements

We thank Dr. Ping Zhao for useful discussions that improved the manuscript. Last, we want to thank Erica Leertouwer for her contribution to the research during her internship.

## Ethics statement

No ethics approval was necessary to conduct this study.

## Contribution to Authorship

JD and SW supervised the project. Study conception and design was executed by HH, JD, RG and SW. HH performed the data gathering with support from JH and AU and analyzed the data with support from JH. Interpretation of results was performed by HH, VG, RG, SW and JD. HH wrote the manuscript. SW and RG obtained the funding for the project. All authors reviewed the manuscript and approved the final version for publication.

