## Supporting Information for "Sotalol dose optimization for fetal tachycardia: a pregnancy physiologically based pharmacokinetic model study"

*[1]Department of Pharmacy, Division of Pharmacology and Toxicology, Radboud University Medical Center, Nijmegen, The Netherlands; [2]Department of Obstetrics and Gynecology, Radboud University Medical Center, Nijmegen, The Netherlands; [3]Department of Intensive Care, Radboud University Medical Center, Nijmegen, The Netherlands [4]Department of Neonatal and Pediatric Intensive Care, Erasmus MC-Sophia Children's Hospital, Rotterdam, The Netherlands*

**Corresponding author –** H. van Hove, Department of Pharmacy, Pharmacology and Toxicology, Radboud university medical center, Geert Grooteplein Zuid 10, 6525 GA Nijmegen, The Netherlands; +3162966931;

### 1. Caco-2 transfer data

The transfer of sotalol over a Caco-2 monolayer was previously reported by Tian et al. [1]. The apparent permeability coefficient ($P_{app}$), which represents the permeability of sotalol over the Caco-2 barrier, was estimated by the following calculation:

$$P_{app} = dQ / (dt * A * C_0)$$

where dQ / dt is the rate of appearance of drugs on the receiver side, A is the surface area of the monolayer and $C_0$ is the initial concentration in the donor compartment. Calculations were conducted for both the experiments in apical-to-basolateral and basolateral-to-apical direction.

The following Papp values were used as input parameters for the placental transfer prediction tool of the Simcyp permeability-limited placenta model:

| | Maternal-to-placental transfer | Placental-to-fetal transfer |
|---|---|---|
| $P_{app}$ (x $10^{-6}$ cm/s) | 0.358 ± 0.006 | 0.422 ± 0.076 |

### 2. Sotalol compound model

Although a sotalol compound model was readily available in the Simcyp repository, the model was not suitable for oral drug dosing predictions. To better estimate absorption of the drug, the advanced dissolution, absorption and metabolism (ADAM) model was used. The input used for the optimised sotalol compound model can be found in Table S1. Simulations were performed using the CVODES option for the ordinary differential equation solver on default Nonstiff mode. For the formulation of the solid drug, immediate release enteric-coated tablet with a triggering pH of 4.9 was used.

**Table S1.** Sotalol dependent input parameters used for the compound model

| 3. | Model parameter / option | Input in Simcyp | User Input / Predicted | Remarks |
|---|---|---|---|---|
| Phys Chem and Blood Binding | MW (g/mol) | 272.36 | [2] / PubChem | |
| | Log $P_{O:W}$ | 0.37 | [2] / PubChem | |
| | Drug type | Ampholyte | User input | |
| | pKa | 8.28/9.72 | | Strongest acid/strongest base pKa |
| | B/P | 1.02 | [2] | |

| | | | | |
|---|---|---|---|---|
| | fu in plasma | 1 | [2] | |
| | Main plasma binding protein | HSA | | |
| Absorption | Absorption model | ADAM | | Intrinsic solubility = 18.414 |
| | $fu_{gut}$ | 0.5575 | Predicted | |
| | $P_{eff}$,man ($10^{-4}$ cm/s) | 1.4 | User input | |
| Distribution | Distribution model | Full PBPK | | |
| | $V_{ss}$ (L/kg) | 1.3538 | Predicted | |
| | Prediction method | Method 2 | | Rodgers and Rowland model |
| Elimination | $CL_R$ (L/h) | 9.9 | [3] | |
| | $CL_{add}$ (L/h) | 0.5 | | |
| Transport | $CL_{PDM}$ (L/h/mL placenta) | 0.030737 | Predicted | Papp values scaled to whole placenta villous surface area |
| | $CL_{PDF}$ (L/h/mL placenta) | 0.03331 | Predicted | |

Abbreviations: advanced dissolution, absorption and metabolism (ADAM), blood-to-plasma partition ratio (B/P), additional systemic clearance ($CL_{add}$), renal clearance ($CL_R$), total placental clearance in maternal-to-placental direction ($CL_{PDM}$), total placental clearance in fetal-to-placental direction ($CL_{PDF}$), unbound fraction in plasma (fu), unbound fraction of drug in enterocytes ($fu_{gut}$), human serum albumin (HSA), molecular weight (MW), neutral species octanol : buffer partition coefficient (Log $P_{O:W}$), human jejunum effective permeability ($P_{eff,man}$), volume of distribution at steady state using tissue volumes for a population representative of healthy volunteers population ($V_{ss}$).

#### 3. Healthy volunteer and pregnancy pharmacokinetic data search queries

The PubMed database was searched for pharmacokinetic data. For the term ‘drugname’ were ‘Sotalol’, ‘beta-Cardone’, ‘Sotalolum’, ‘DL-Sotalol’ or ‘3930-20-9’ used, respectively. Titles and abstracts of all search results were screened to check if actual PK data were provided in the publication. To extend our search strategy, the ‘Similar articles’ overview and references were checked to identify relevant articles which were missed with the initial search queries. Eventually, seven articles were included for the healthy volunteer population and three studies for the pregnant population. The pharmacokinetic data of the studies of the healthy volunteer population is displayed in Table S2 and of the pregnant population in Table S3.

<u>Search query for healthy volunteer pharmacokinetic studies:</u>

((Pharmacokinet*[Title/Abstract]) AND (***DRUGNAME****[Title]) AND (Healthy[Title] OR volunteer*[Title] OR adult*[Title] OR subject*[Title] OR Man[Title] OR Men[Title] OR Woman[Title] OR Women[Title] OR male*[Title] OR female*[Title])) OR (("Pharmacokinetics of"[Title]) AND ("***DRUGNAME***"[Title]) OR (“***DRUGNAME*** pharmacokinetics”[Title]))

Sotalol → 64 results (17-10-2023)

<u>Search query for pregnancy pharmacokinetic studies:</u>

(***DRUGNAME**** [Tiab]) AND ((Pharmacokinetic*[Tiab] OR kinetic*[Tiab]) OR (‘Plasma concentration’[Tiab] OR ‘Plasma level’[Tiab] OR (***DRUGNAME*** / Pharmacokinetics[MeSH] OR ***DRUGNAME*** / blood[MeSH]))) AND (Pregnancy [MeSH] OR Pregnancy*[Tiab] OR Pregnant*[Tiab])

AND (Fetus [MeSH] OR Foetus*[Tiab] OR Fetus*[Tiab] OR fetal*[Tiab] OR Foetal*[Tiab] OR Embryo*[Tiab] OR (Fetal blood [MeSH] OR ‘Cord blood’[Tiab] OR ‘Umbilical vein’[Tiab] OR ‘Umbilical artery’[Tiab]) OR (Maternal-fetal exchange[MeSH] OR ‘Transplacental Exposure’[Tiab]))

Sotalol → 11 results (17-10-2023)

**Table S2.** An overview per administration route of the pharmacokinetic studies of healthy volunteers suitable to verify the PBPK model

| Study | N | Dose | Administration time | Age range | Duration (hours) | Proportion of females | Ref |
|---|---|---|---|---|---|---|---|
| **Intravenous** | | | | | | | |
| Somberg et al. 2010 | 15 | 75 mg | 2.5 h | 19-45 | 48 | 0.60 | 4 |
| Salazar et al. 1997 | 12 | 0.5 mg/kg<br>1.5 mg/kg<br>3.0 mg/kg | 2 min | 28-38 | 72 | 1.00 | 5 |
| Poirier et al. 1989 | NS | 0.25 mg/kg<br>0.50 mg/kg<br>1.00 mg/kg<br>2.00 mg/kg | 5 min | 22-25 | 48 | 0.0 | 6 |
| **Oral** | | | | | | | |
| Darpo et al. 2014 | 11 | 160 mg | | 18-45 | 22.5 | 1.0 | 7 |
| Démolis et al. 2005 | 25 | 160 mg | | 19-36 | 24 | 1.0 | 8 |
| Hanyok et al. 1993 | 18 | 160 mg | | NS | 36 | NS | 9 |
| Sundquist et al. 1979 | 12 | 160 mg | | 25-57 | 32 | NS | 10 |

Abbreviations: Not specified (NS), number (N), milligram (mg), milligram per kilogram bodyweight (mg/kg), minute (min), hour (h), reference (Ref).

Poirier et al., Démolis et al. and Hanyok et al. Were previously used to determine the renal CL, so can indirectly be regarded as articles used for model development.

**Table S3.** An overview per administration route of the pharmacokinetic studies of pregnant women suitable to verify the pregnancy PBPK model

| Study | N | Dose | Administration time | Age range | Duration (hours) | GA (weeks) | Ref |
|---|---|---|---|---|---|---|---|
| **IV** | | | | | | | |
| O'Hare et al. 1983 | 6 | 100 mg | 5 min | NS | 24 | 32-36 | [11] |
| **PO** | | | | | | | |
| O'Hare et al. 1983 | 6 | 400 mg | | NS | 36 | 32-36 | [11] |
| Erkkola et al. 1982 | 16 | 80 mg | | 25-34 | 3 | 37-40 | [12] |
| Starodubsteva et al. 2023 | 30 | 80-240 mg BID | | 28-34 | Range, max 1400 | 30-40 | [13] |

Abbreviations: Gestational age (GA), not specified (NS), number (N), milligram (mg), twice daily (BID), reference (Ref).

### 4. Healthy volunteer PBPK model verification

Visual predictive checks for sotalol IV administration are shown in Figure S1 for healthy volunteers. Outcomes of a single oral administration are depicted in Figure S2. Overall, the predictions were considered acceptable to continue simulating sotalol plasma concentrations during pregnancy.

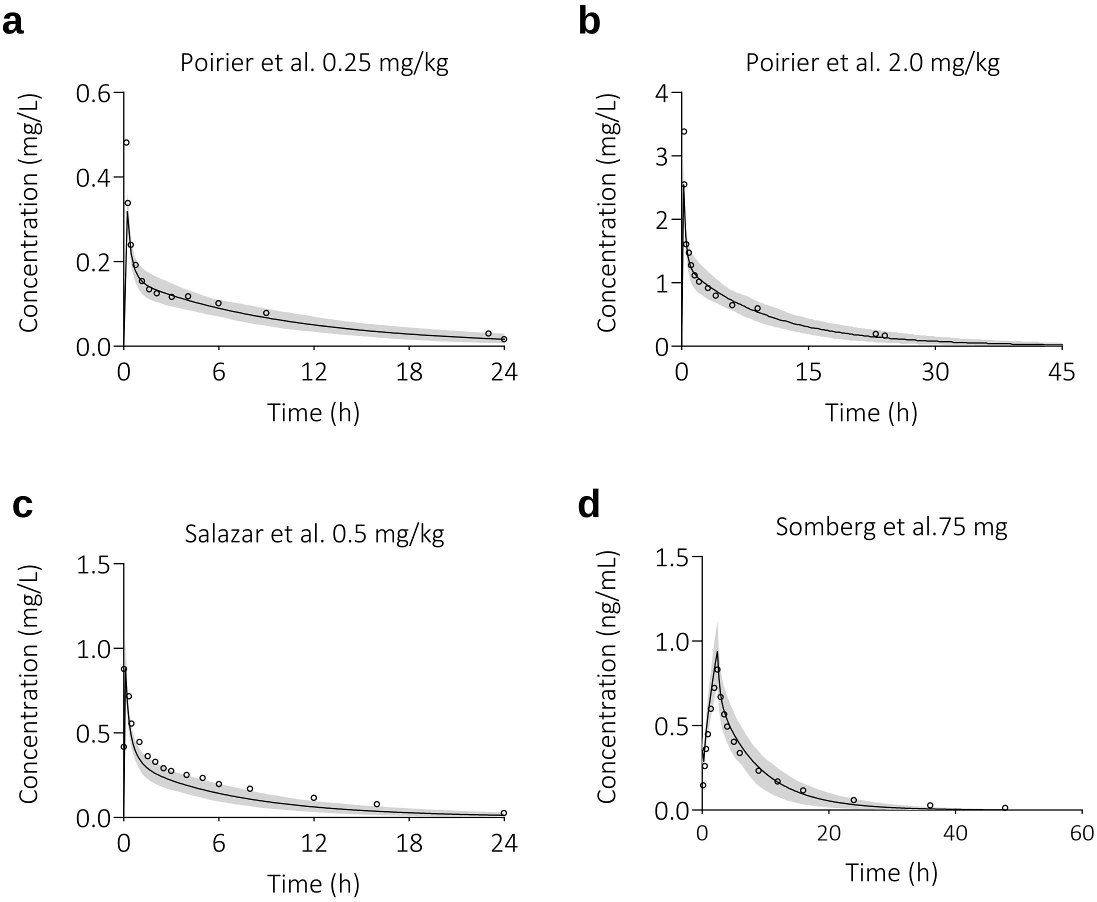


**Figure S1. Prediction of sotalol plasma concentration-time profiles in healthy volunteers after single intravenous (IV) administration.** The solid line is the predicted mean of the simulated population and the shaded area represents the 5$^{th}$ to 95$^{th}$ percentile of the virtual population. Open circles are the observed data (a & b [6], c [5] and d [4]). Abbreviations: milligram (mg), milligram per kilogram bodyweight (mg/kg), hour (h).

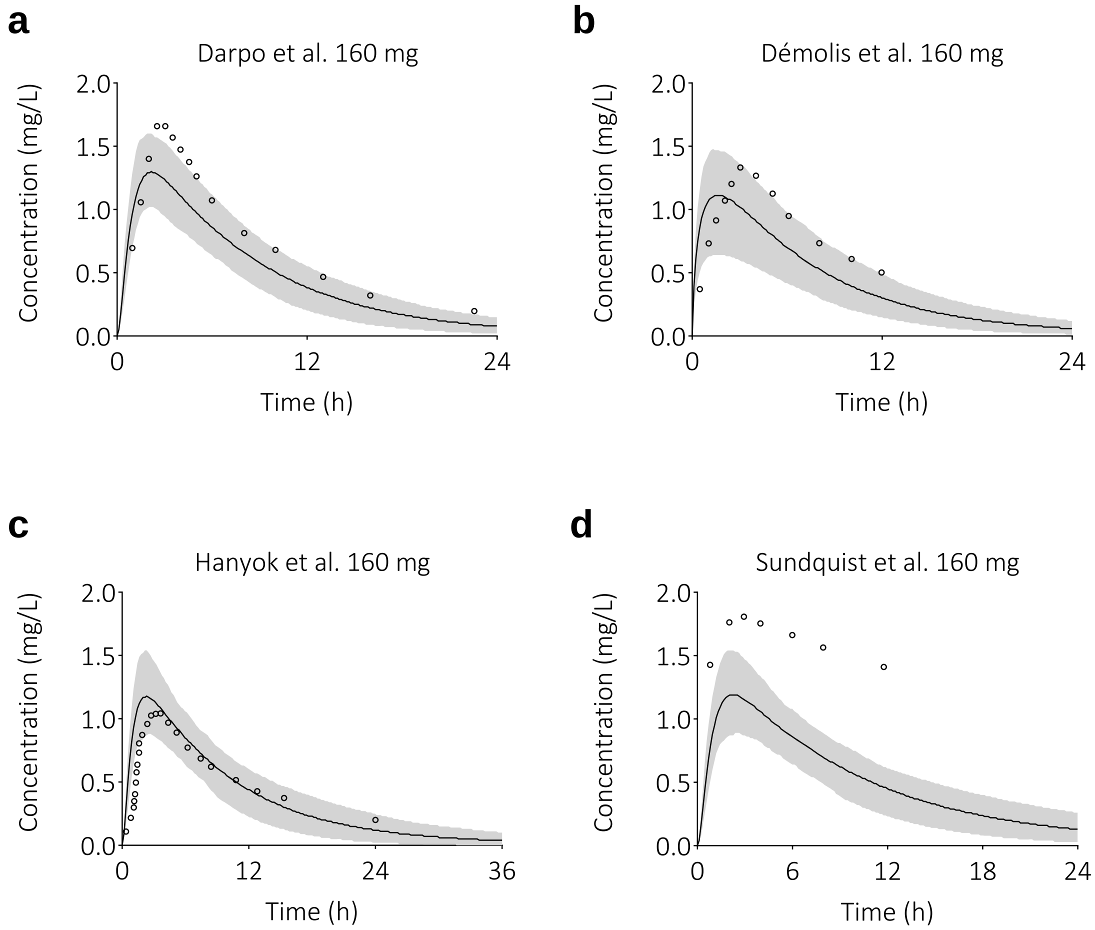


**Figure S2. Prediction of sotalol plasma concentration-time profiles in healthy volunteers after single oral (PO) administration.** The solid line is the predicted mean of the simulated population and the shaded area represents the 5$^{th}$ to 95$^{th}$ percentile of the virtual population. Open circles are the observed data (a [7], b [8], c [9] and d [10]). Abbreviations: milligram (mg), hour (h).

### 5. Pregnancy PBPK model verification

Visual predictive check for a single oral administration is shown in Figure S3 for pregnant women. Maternal visual predictive checks for multiple oral administrations are depicted in Figure S4, while fetal predictive checks are presented in Figure S5. Simulations that exceeded a study duration of 7 days were conducted using the redefine subjects over time option (daily redefinition). Overall, the predictions were considered acceptable to continue simulating current dosing regimen to enable dose re-evaluation.

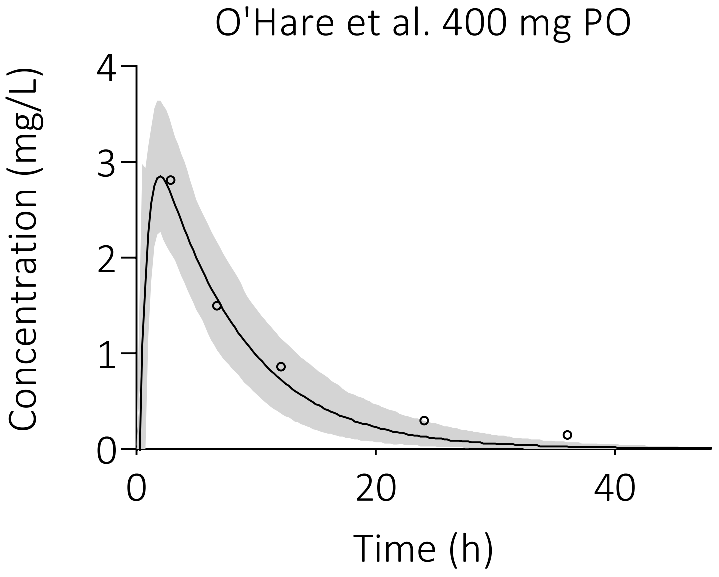


**Figure S3. Prediction of a sotalol plasma concentration-time profile in pregnant women after a single oral (PO) administration.** The solid line is the predicted mean of the simulated population and the shaded area represents the 5th to 95th percentile of the virtual population. Open circles are the observed data [11]. Abbreviations: milligram (mg), hour (h).

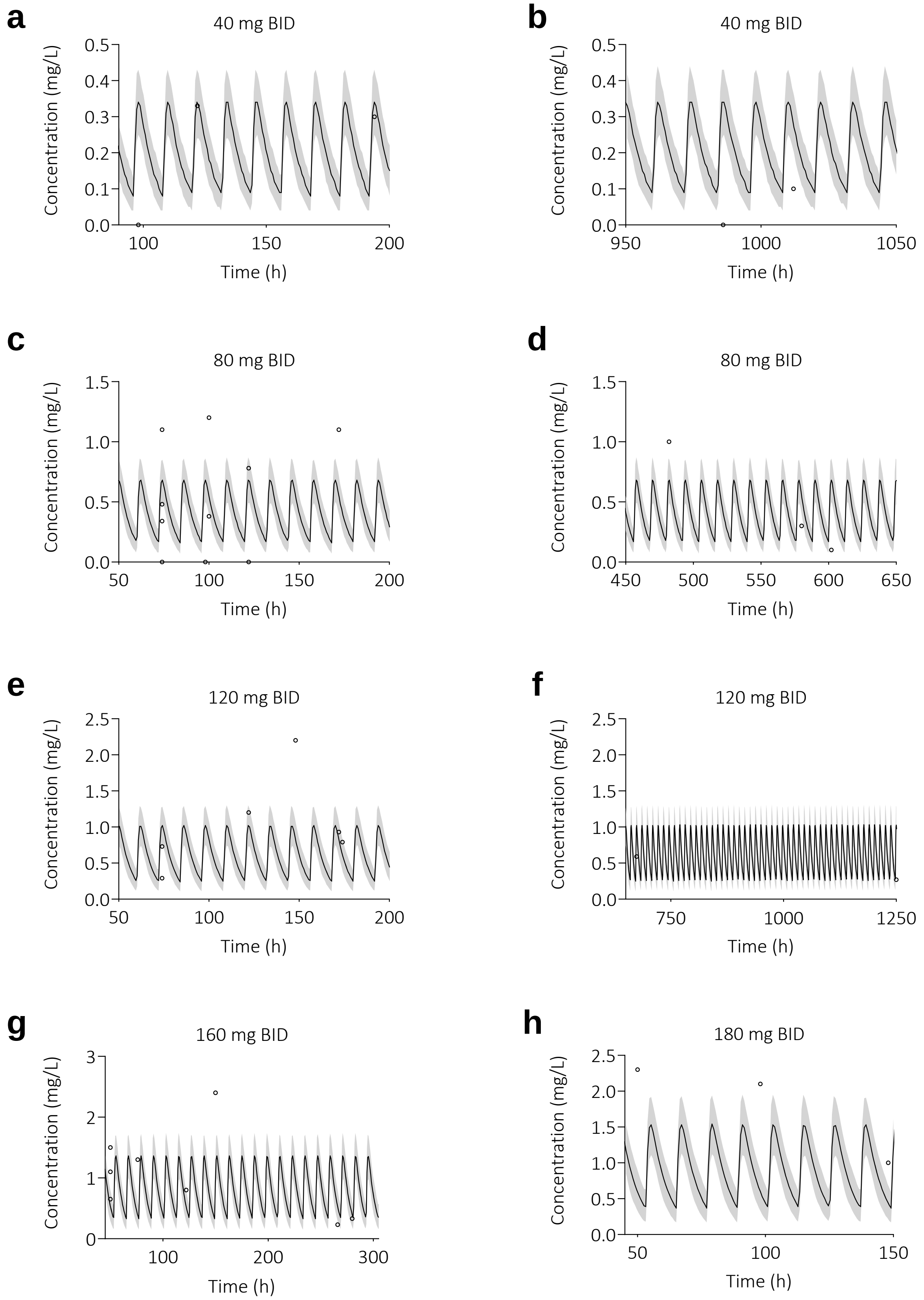
a
40 mg BID
b
40 mg BID
c
80 mg BID
d
80 mg BID
e
120 mg BID
f
120 mg BID
g
160 mg BID
h
180 mg BID
Concentration (mg/L)
Time (h)

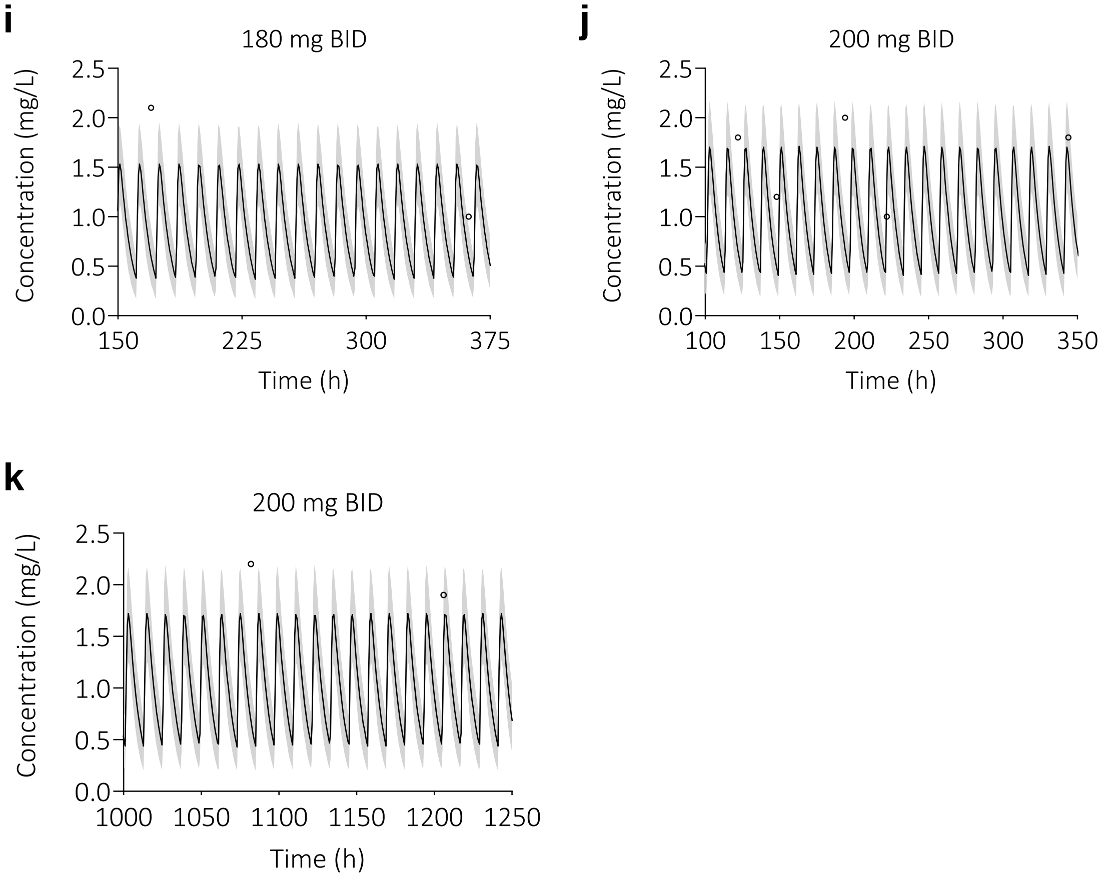


**Figure S4. Prediction of sotalol plasma concentration-time profiles in pregnant women after multiple oral administrations.** The solid line is the predicted mean of the simulated population and the shaded area represents the 5$^{th}$ to 95$^{th}$ percentile of the virtual population. Dosing regimens of 80 (a & b), 160 (c & d), 240 (e & f), 320 (g & h), 360 (i) and 400 mg/day (j & k) were simulated. Open circles are the observed data [13]. Abbreviations: twice daily (BID), milligram (mg), hour (h).

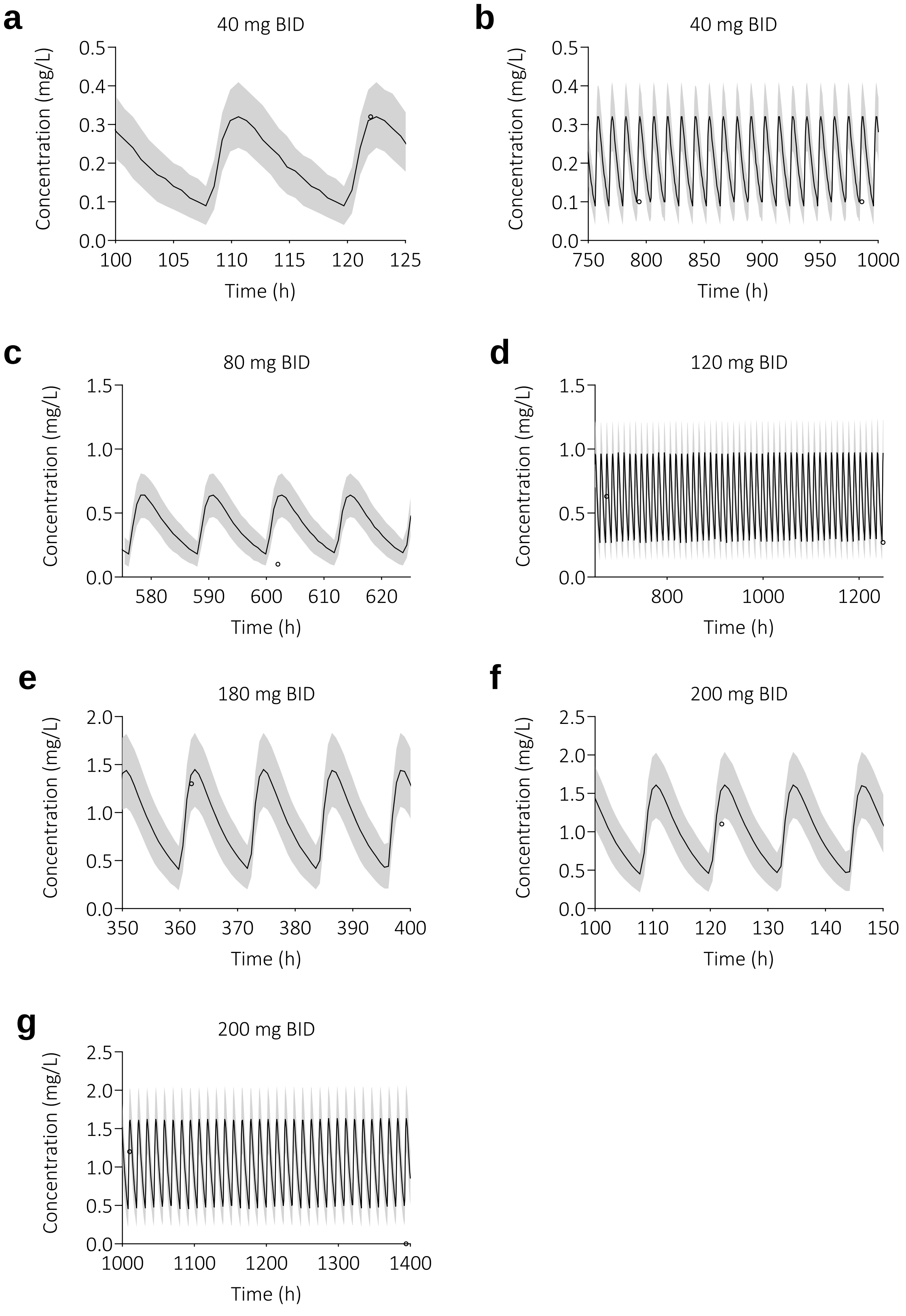


**Figure S5. Prediction of sotalol plasma concentration-time profiles in unborn children after multiple maternal oral administrations.** The solid line is the predicted mean of the simulated population and the shaded area represents the $5^{th}$ to $95^{th}$ percentile of the virtual population. Dosing regimens of 80 (a & b), 160 (c), 240 (d), 360 (e) and 400 mg/day (f & g) were simulated. Open circles are the observed data [13]. Abbreviations: twice daily (BID), milligram (mg), hour (h).

### 6. Current dosing regimen

To enable re-evaluation of current dosing guideline, maternal (Figure S6) and fetal (Figure S7) exposure during sotalol therapy was simulated. Each dose was given orally twice daily (e.g., 12 hours apart) in a fasted state, and therapy was continued for 72 hours. Hundred virtual subjects were simulated, with a gestational age of 32 weeks. The starting dose was 160 mg/day, which was increased in steps of 80 mg/day to a maximum of 480 mg/day. The maternal toxic level of 2.5 mg/L and the fetal therapeutic $C_{trough}$ concentration window of 0.4-1.0 mg/L were used as reference values to compare predicted exposures to [14].

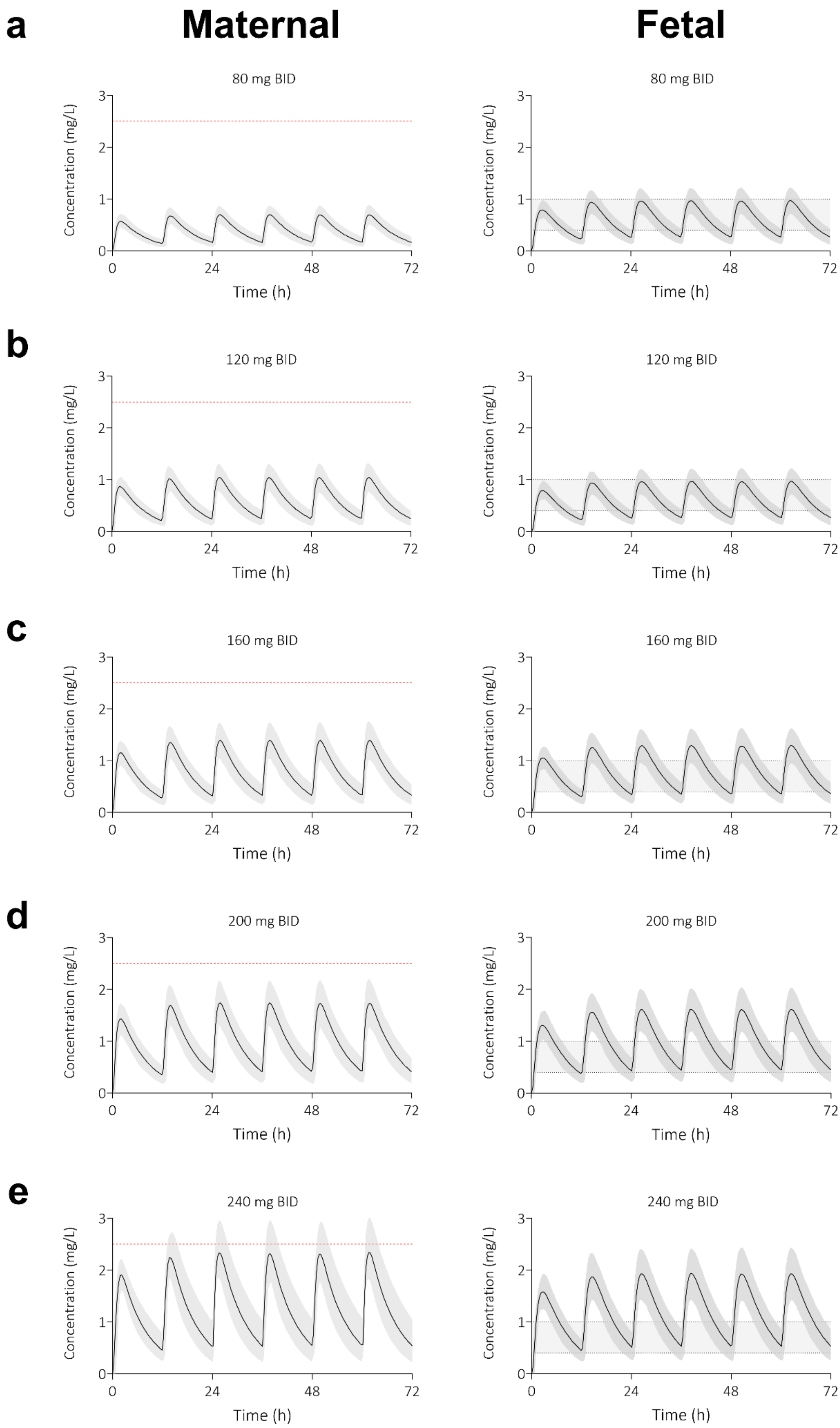

a
Maternal
Fetal
80 mg BID
b
120 mg BID
c
160 mg BID
d
200 mg BID
e
240 mg BID
Concentration (mg/L)
Time (h)

**Figure S6. Predicted plasma concentration-time profiles of sotalol for pregnant women and their unborn children after multiple oral doses.** Simulation duration was 72 hours. The shaded area represents the 5th to 95th percentile of the virtual populations, the red dotted line the maternal toxic level of 2.5 mg/L, and the grey area the fetal therapeutic $C_{trough}$ window of 0.4–1.0 mg/L. Dosing scenarios of 160 (a), 240 (b), 320 (c), 400 (d) and 480 mg/day (e) were simulated, respectively. Abbreviations: oral (PO), milligram (mg), twice daily (BID).

### 7. Prospective simulations

To assess whether sotalol plasma exposure in mother and child could be optimised, simulations for three times a day oral prescription were executed. In Figure S7 is shown that predicted plasma concentrations for the mother remain below the toxic level of 2.5 mg/L upon 40 mg TID, however, fetal plasma concentrations are insufficient for successful therapy.

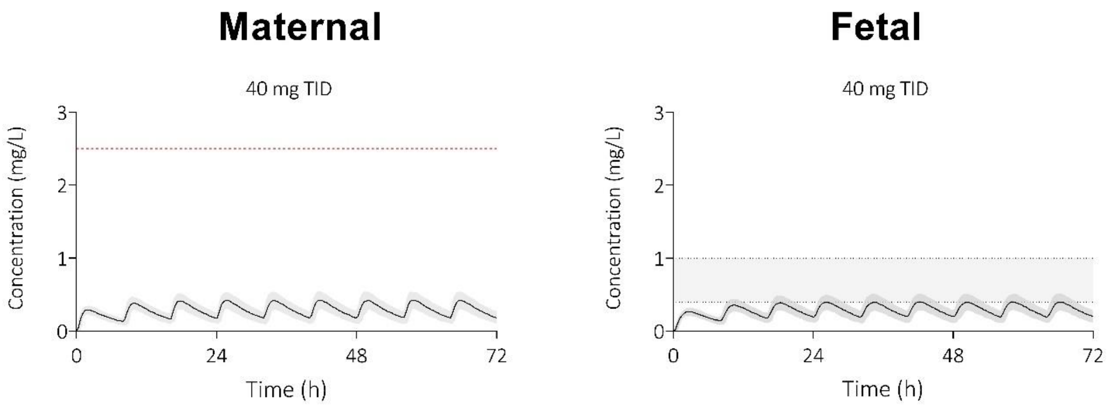


**Figure S7. Prospective simulations of sotalol plasma concentration-time profiles for pregnant women and their unborn children after an oral dose of 120 mg/day.** Simulation duration was 72 hours. The shaded area represents the 5th to 95th percentile of the virtual populations, the red dotted line the toxic level of 2.5 mg/L and the grey area the fetal therapeutic $C_{trough}$ window of 0.4-1.0 mg/L (B&D). Abbreviations: three times a day (TID), milligram (mg).
